# Dengue virus transmission in Italy: surveillance and epidemiological trends up to 2023

**DOI:** 10.1101/2023.12.19.23300208

**Authors:** Francesco Branda, Taishi Nakase, Antonello Maruotti, Fabio Scarpa, Alessandra Ciccozzi, Chiara Romano, Simone Peletto, Ana Maria Bispo de Filippis, Luiz Carlos Junior Alcantara, Alessandro Marcello, Massimo Ciccozzi, José Lourenço, Marta Giovanetti

## Abstract

Dengue virus circulation is on the rise globally with increased epidemic activity in previously unaffected countries, including within the European region. In 2023, global dengue activity peaked, and Italy reported the highest number of dengue cases and local chains of transmission to date. By curating several sources of information, we introduce a novel data repository focused on dengue reporting in Italy. We integrate geographic, epidemiological, genomic and climatic spatio-temporal data to present an overview of transmission patterns of the past eight years related to circulating viral lineages, geographic distribution, hotspots of reporting, and the theoretical contribution of local climate. This study contributes to a better understanding of the evolving scenario in Italy, with the potential to inform reassessment and planning of adequate national and European public health strategies to manage the emergence of dengue.

## Introduction

Dengue fever is a mosquito-borne viral infection caused by the Dengue virus (DENV), which belongs to the *Flaviviridae* family. Common symptoms include high fever, intense headaches, and significant muscle pain [1]. Transmission primarily occurs through the bites of *Aedes spp.* infected mosquitoes [1]. In recent years, there has been a significant global increase in incidence, leading to impacts on and concerns for public health systems [2]. One of the primary drivers of this surge is climate change [3,4]. Ongoing climatic changes, including temperature fluctuations and altered rainfall patterns, have not only contributed to the increased incidence but also facilitated geographic spread, notably towards higher altitudes [3]. Historically, Europe has experienced mostly imported cases of dengue, with sporadic autochthonous transmission [2]. However, autochthonous cases have been surging in the past five years, with significant reports from Spain [5], Croazia [6], France [7] and Italy [8]. In 2023, Italy reported its highest ever number of locally transmitted dengue [9].

In the present study, we integrate several sources of geographic, genetic, epidemiological and climate data, presenting a comprehensive overview of the spatiotemporal patterns and possible drivers of dengue activity in Italy.

## Methods

To assess the trends of DENV in Italy, we collected epidemiological data from the weekly reported cases published by the Istituto Superiore di Sanità (ISS) on the EpiCentro website, which can be accessed at [9]. The process started with downloading and pre-processing all bulletins from the EpiCentro website into CSV files. These are now stored in a publicly accessible GitHub repository, which is regularly updated with new Italian data as they become available. The repository can be found at https://github.com/fbranda/arbovirus, and a detailed synopsis of the file structure is provided in **Table S1**.

### Reporting risk model

We modeled reported counts per area i as *Y*_*i*_, (i = 1 to n) using a Poisson distribution with mean *E*_*i*_× *θ*_*i*_, where *E*_*i*_ is the expected counts and *θ*_*i*_ is the relative risk in area i. Then, the log risks are modeled with a sum of an intercept to model the overall disease risk level and random effects that account for extra-Poisson variability in the observed data [10]. Areas with relative risks *θ*_*i*_ > 1 and *θ*_*i*_ < 1 are areas with high and low risks, respectively. Areas with *θ*_*i*_ = 1 has the same risk as expected from the standard population. In this work, the model for disease mapping is expressed as:

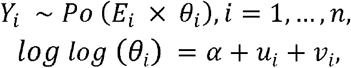

where a denotes the overall risk level, *u*_*i*_ is a spatial structured random effect that models the spatial dependence between the relative risks, and *v*_*i*_ is an unstructured exchangeable random effect that models uncorrelated noise.

### Detection of clusters

To identify spatio-temporal clusters (i.e., geographic areas in which the number of observed incidence events is significantly different from that predicted by a random distribution) we used Kulldorff’s scan statistics [11]. This method uses a moving window, which can take various shapes, such as circles or ellipses, and different sizes, running systematically over the entire study area. For each position and size of the window, the scan statistic calculates a probability index based on the ratio of the number of events observed within the window to the number of events expected under a null hypothesis of random distribution (i.e. the events are randomly distributed across the entire study area). The Kulldorff’s scan statistics then compares the actual data collected with this random distribution for each window, and calculates the likelihood ratio (LR) to determine whether the observed cluster is statistically significant or could be due to chance:

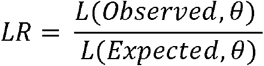

where L(*Observed, θ*) represents the likelihood of the observed data within the window, considering the parameters *θ* of the alternative hypothesis, and L(*Expected, θ*) represents the likelihood of the expected data within the window, also considering the parameters *θ* of the alternative hypothesis. An LR greater than 1 indicates that there are more observed cases within the window than expected.

To determine the statistical significance of a cluster, LR is compared to a threshold value. This threshold is often represented as a -log(p-value). The p-value quantifies the probability of observing a cluster as extreme as, or more extreme than, the one identified under the null hypothesis that cases are randomly distributed. A lower p-value indicates a more significant cluster. More specifically, the -log(p-value) is compared to the critical values from a chi-squared distribution (χ^2^). If the log(p-value) is greater than or equal to the critical value from the chi-squared distribution at a specified significance level (often set at 0.05 or 0.01), the cluster is deemed statistically significant, and the null hypothesis of random distribution within the spatial window is rejected. The outcome of this significance calculation guides the interpretation of the spatial window or cluster. A statistically significant cluster suggests that the observed pattern of events is unlikely to be the result of random chance and may indicate the presence of a true cluster. In contrast, a non-significant cluster implies that the observed pattern can be reasonably explained by random variation. All analyses were performed using R (v4.3.1) [4] and we used SaTScan software (v10.1.2) (https://www.satscan.org/) [5] to perform geographic disease surveillance for early detection of disease outbreaks and evaluate the statistical significance of disease cluster alarms.

### Dated phylogenetics

We analyzed all available genome sequences from GISAID (DENV 1 n=7, DENV 3 n=3) along with reference strains (n=95 for DENV1 and n=140 for DENV3 respectively). These sequences were aligned using MAFFT [12] and manually refined in Aliview [13] to eliminate anomalies. The GTR model, deemed the best fit by ModelFinder in IQ-TREE2 [14], was employed for the initial estimation of maximum likelihood phylogenetic trees, with robustness validated through 1,000 bootstrap replicates. TempEst [15] aided in detecting temporal signals, while BEAST facilitated the inference of time-scaled phylogenetic trees. A comprehensive model selection process, incorporating path-sampling and steppingstone methods, identified the uncorrelated relaxed molecular clock model, utilizing the SRD06 model and Bayesian Skygrid coalescent model, as optimal for Bayesian analysis [15,16]. We conducted phylogeographic analysis to map the virus’ spatial diffusion, using discrete sampling locations (countries) and an asymmetric model of location transition with BSSVS [17]. To ensure thoroughness, MCMC runs were performed in duplicate over 100 million iterations, achieving an effective sample size (ESS) of over 200. The final maximum clade trees, post 10% burn-in exclusion, were compiled using TreeAnnotator and displayed in FigTree v1.4.4.

### Analysis of long-term temperature trends

Long-term trends in local temperature were estimated using a linear regression model on each geographical pixel of monthly satellite temperature time series from 1979 (January) to 2023 (September) [18]. In the main text we report the estimated slopes (all had p-value< 0.0001).

### Climate-based transmission suitability

We estimated theoretical, dengue virus climate-based transmission suitability, as per Nakase et al [3] for each geographical pixel available in satellite climate data from Copernicus.eu [18]. This suitability measure (index P) measures the reproductive, transmission potential of a single adult female mosquito during its lifetime in a fully susceptible host population [19]. Since satellite data was only available up to September 2023, all outputs in the main text related to transmission suitability were summarized per year using climate data between January and September of each year.

## Results

### Spatio-temporal Dynamics of Dengue in Italy

The spatio-temporal dynamics of reported dengue in Italy are summarized in **Figure 1**. Between 2015 and 20 November 2023, there have been a total of 1028 reported dengue cases nationally, including local and imported cases. Of these 253 were reported between 2015 and 2017, for which there was no available geolocation (**Figure 1AB**). Between 2018 and 2023, a total of 775 cases were reported, with the Northeast, Northwest and Center macroregions each experiencing about 33% of cases (N=269, 256, 225, respectively), and the South/Insular experiencing only ∼3% of cases (N=25).

**Figure 1.**
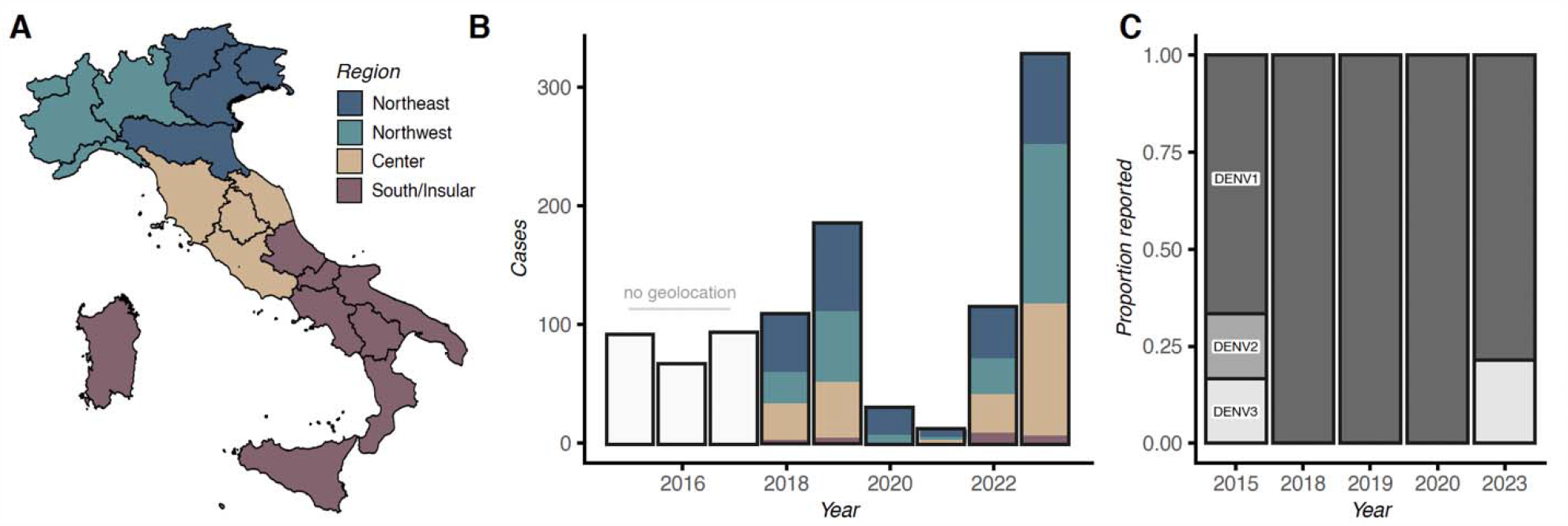
Spatiotemporal Dynamics of Dengue in Italy. **A)** Map of Italy with regions (black boundaries) and macroregions (color legend). **B)** Time series of reported dengue cases. Reports between 2015-2017 had no geolocation, while those after 2018 are aggregated by macroregion. **C)** Yearly count of available DENV genome sequences by serotype in the GISAID repository (January 2015 to November 20, 2023). Missing years had no genomes.

During 2023, the year with highest ever reported cases (N=327, ∼31%), there were 75 cases in the Northeast, 134 in the Northwest, 111 in the Center and only 7 in the South/Insular macroregions (**Figure 1B**). Among these, 82 were confirmed as autochthonous and attributable to four distinct chains of transmission within the regions of Lodi in the Northwest, and Latina, Rome (including metropolitan areas), Anzio, all in the Center macroregion.

Genomic data, derived from the complete genome sequences available in the public repository (GISAID), revealed that three dengue serotypes have circulated in Italy in the last eight years (**Figure 1C**). In 2015, DENV1, DENV2, and DENV3 were reported as circulating, with DENV1 being the predominant serotype. Between 2018 and 2020, DENV1 was the only serotype present in the database, while in 2023 both DENV1 and DENV3 were detected. This recent trend in Italy aligns with the global trends observed since early 2023, where there has been an increase in the circulation of DENV1 and DENV3, particularly across Latin American countries [20, 21].

### The 2023 Epidemiological Dynamics of Dengue in Italy

By November 20, 2023, active dengue virus transmission was reported in 15 of Italy’s 21 regions. This wide geographical range has prompted a nationwide movement, aimed at comprehensively assessing and identifying gaps in the public health response and preparedness. **Figure 2A** delineates regions with high Dengue incidence in 2023, categorized based on weekly case reports: Zero/Low includes regions like Abruzzo and Aosta Valley with up to five cases; Medium covers regions from Campania to Veneto with six to fifteen cases; and High represents Emilia Romagna, Lazio, Lombardy, and Piedmont, reporting over fifteen cases. Notably, Lombardy and Lazio represent 33% and 25% of the total cases, respectively, highlighting significant reporting concentrations.

**Figure 2.**
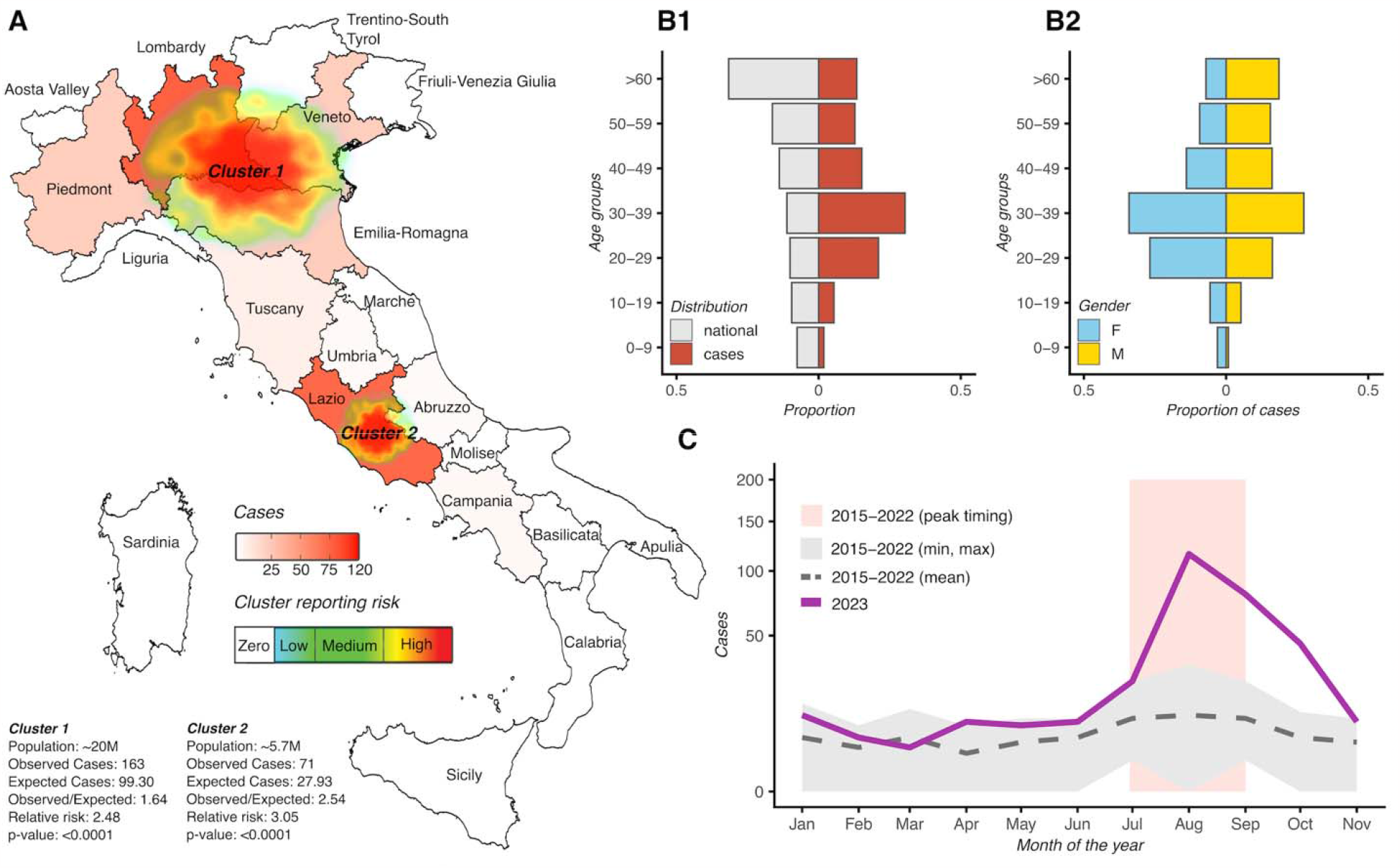
The 2023 Epidemiological Dynamics of Dengue in Italy. **A)** Identification of reporting hotspots across Italian regions in 2023, including a summary of key characteristics within the identified clusters. **B1)** Breakdown of reported cases by age group. In red the proportion of cases per age group, and in grey the proportion of the national population per age group. **B2)** Breakdown of reported cases by gender. In blue the proportion of cases reported as female per age group, and in yellow the proportion of cases reported as male per age group. For the period 2015-2022: grey dashed line is the monthly reporting mean; grey area marks the limits of the minimum and maximum reported cases over the months; pink area marks the typical time window of peak reporting. For 2023, the full purple line is the total cases per month. The y-axis is square-root transformed for visualization.

Two major clusters were identified: the first extended across a diagonal axis within the Northwest and Northeast macroregions, from Piedmont to Emilia Romagna, covering over 20 million people; the second was located in Lazio, with a population of 5 million. The relative risk (RR) of dengue infection in these clusters was 2.48 in the first and 3.05 in the second. The median age of individuals was 37 years, with a slight male predominance of 51.68% (**Figure 2B**). In general, cases were more frequently reported among the 20-29 and 30-39 age groups, contrasting with the national age profile of the population (**Figure 2B1**). Between genders, case were also concentrated among the 20-29 and 30-39 age groups for females, but were more uniformly distributed for males older than 20 years (**Figure 2B2**). Compared to the monthly average trends in reported cases with the period 2015-2022, the year 2023 was different in both seasonal dynamics and size (**Figure 2C**). Specifically, reports for 2023 were consistently higher than the historical average across the months (with the exception of March). Peak reporting in 2023 was in August (N=116) well within the historical time window of peak reporting, but peaking much higher (ratio = 9.6) and only approaching average historical levels late in November.

### Phylodynamic Analysis of co-circulating DENV1 and DENV3 in 2023

We combined all available partial and complete DENV genome sequences from Italy, primarily generated during the 2023 epidemic, with other DENV1 and DENV3 genomes from public repositories. Our phylogenetic analysis revealed that the novel 2023 isolates belonged to DENV1 genotype V (DENV1-V) and DENV3 genotype III (DENV3-III). These are currently the predominant genotypes circulating in Latin American countries [20, 21].

Further analysis of DENV1-V indicated two major introductions into Italy, estimated to have occurred in early and late December 2022 (High Posterior Density interval ranging from July 24 2022, to June 10 2023) (**Figure 3A**). Discrete phylogeographic reconstruction also suggested that South America, primarily represented by Brazilian sequences, was a central hub for multiple viral introductions into Italy (**Figure 3B**).

**Figure 3.**
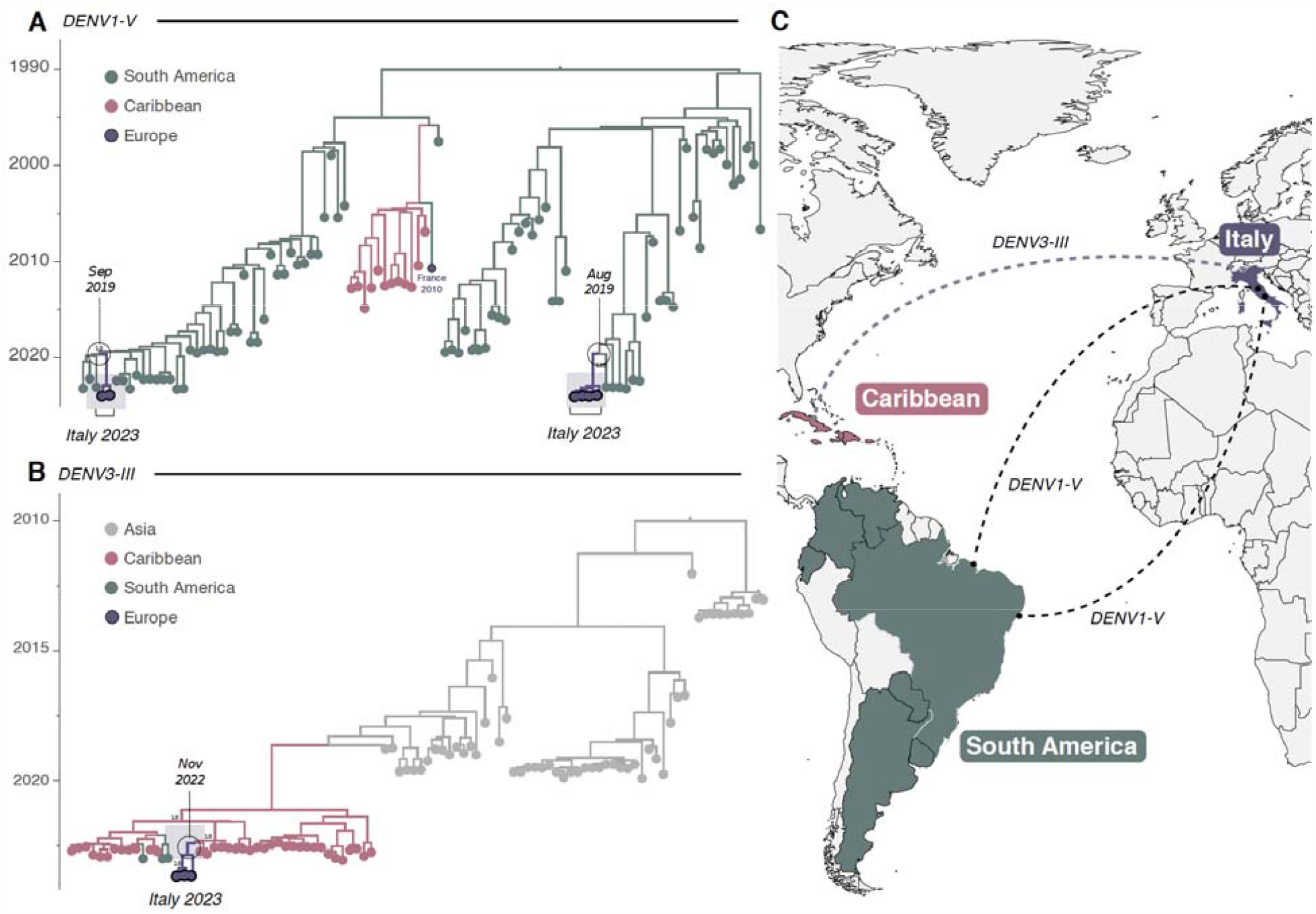
Phylodynamic analysis of DENV1-V and DENV3-III in Italy. **A)** Time-scaled phylogeographic tree of DENV1-V (including all Italian strains currently available *n*=7 plus *n*=95 GenBank sequences). Colors represent different sampling locations. **B)** Time-scaled phylogeographic tree of DENV3-III (including all Italian strains currently available *n*=3 plus *n*=140 GenBank sequences). Colors represent different sampling locations; **C)** Ma displaying the countries involved, with virus transmission paths marked by counter-clockwise lines colore according to the viral source. DENV1-V is shown with a black dashed line, and DENV3-III is represented by a light green dashed line.

We further traced the introduction of DENV3-III to late November 2022 (High Posterior Density interval ranging from August 22 2022, to June 28 2023) (**Figure 3C**). Our findings suggested that the Caribbean (including Cuba), which saw a rise in DENV3-III cases since 2022 [20], might have been the source of this introduction (**Figure 3D**).

### Climate-based Transmission Suitability of Dengue in Italy

We next explored whether climate, an established driver of arboviral transmission potential [3, 4], could shed light on patterns of Dengue virus reporting across Italy. Using climate satellite data, we estimated the theoretical transmission suitability of dengue between 2015 and 2023. Suitability was found to vary across space (**Figure 4A1**) and time (**Figure 4A2**). The far north and the latitude-longitude diagonal across the country, rich in high altitude land, consistently presented the lowest suitability. In contrast, the island of Sicily, the continental coasts and some areas of the northern macroregions consistently presented intermediate-to-high suitability. Notably, these areas also showed high suitability (suitability > 1) for a few consecutive months of the year (**Figure 4B1**). A suitability of 1 is theoretically relevant because it marks the threshold above which a single infected female mosquito could transmit the virus to more than one host during their lifetime.

**Figure 4.**
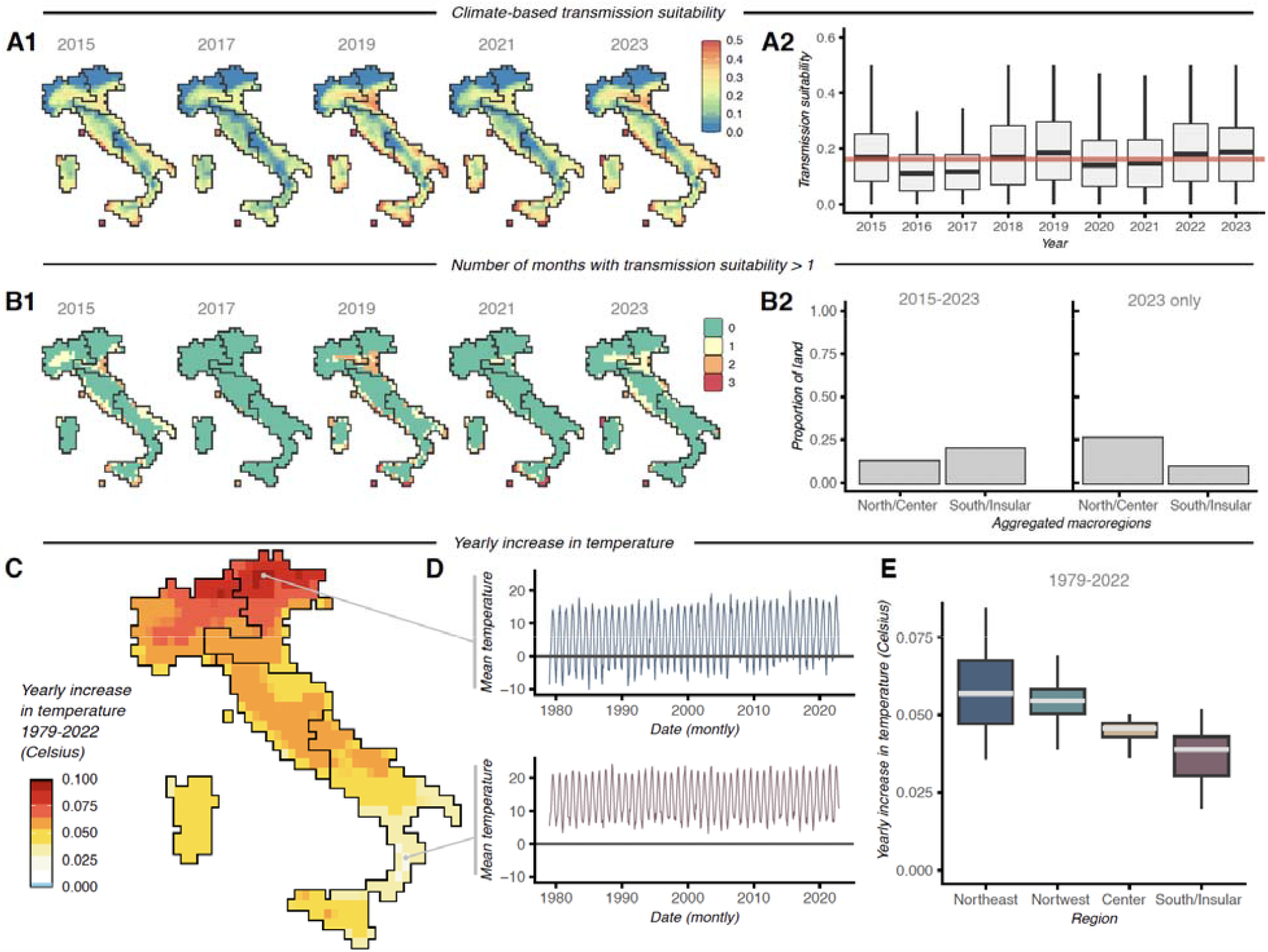
Climate-based Suitability of Dengue in Italy. **A1)** Yearly mean climate-based transmission suitability (index P) per geographical pixel of the climate satellite data; presented are a subset of years. **A2)** Distributions of suitability values per year. The light red horizontal line marks the historical mean (2015-2023). **B1)** Yearly number of months in which climate-base transmission suitability (index P) per geographical pixel of the climate satellite data was above 1; presented are a subset of years. **B2)** Proportion of land (of geographical pixels available in climate data) that present suitability above 1 for at least 1 month of the year. Presented are the historical period (2015-2023) and 2023 only. North/Center is the aggregation of the Northwest, Northeast and Center macroregions. **C)** Map of Italy with estimates of rate of increase (linear slope) in mean yearly temperature (color legend). Black boundaries are macroregions. **D)** Time series of mean monthly temperature (1979-2022) for two geolocations as marked on panel D. Line coloring follows the corresponding macroregion. **E)** Distributions of rates of increase (linear slopes) for each of the macroregions (as presented in panel D).

The year 2023, in which an exceptionally high number of autochthonous cases were reported, presented transmission suitability only slightly above the historical average (**Figure 4B1**). However, 19% of total land in Italy showed suitability above 1 for at least one month in 2023. - In contrast to historical trends but following the geographical distribution of reported autochthonous cases, suitability above 1 occurred mostly in the Northwest, Northeast and Center macroregions. (**Figure 4B2**).

Finally, we quantified historical trends in local climate by estimating rates of change in temperature. Our estimates showed a universal, yearly increase in mean temperature across Italy (**Figure 4C**). The highest rates of increase were found in the Northeast macroregion (**Figure 4D**), followed by the Northwest, Center and South/Insular macroregions (**Figure 4DE**).

## Discussion

This study summarizes reported dengue infections in Italy of the last eight years, introducing a new data repository of incidence reports into the future. The repository is designed to improve accessibility to information by the research and education communities, thereby supporting the generation of knowledge and awareness related to the emergence of dengue in the country.

We explored the potential contribution of local climate to the transmission potential of dengue across Italy by estimating a commonly used climate-based suitability measure. We found that suitability is spatially heterogeneous and generally much lower than that estimated for endemic countries such as Brazil or the Dominican Republic (see e.g. [3,22]). Although not directly quantified, suitability was generally lower at higher altitudes. Reasonable suitability was found along continental coastal areas, the island of Sicily and interior regions of the Northern macroregions. Coincidentally, with the exception of Sicily, these areas were involved in the exceptional emergence of chains of transmission during 2023. However, our analyses also reveal that the majority of land area rarely shows high suitability, with less than 20% with high suitability for at least one month of the year. This suggests weak potential for long-term transmission and sheds light the potential current role of local climate in limiting persistence. Nonetheless, we also uncover that mean yearly temperature has been increasing rapidly, especially in the Northern macroregions, conspicuously matching the areas where West Nile virus and Usutu virus have emerged as public health threats in recent years [23, 24]. While transmission suitability for dengue does not seem to have increased at a similar pace, the effect of rising temperatures is likely to have positive effects on the life cycle and reproduction of mosquito-species generally involved in arboviral transmission. Together, these results underline the need for adaptable mosquito control policies due to environmental changes.

We note historical asymmetries in dengue reporting across Italy. Over the years, a highly variable number of dengue cases have been reported countrywise, with specific relevance regarding higher latitudes (including the official Northwest, Northeast and Center macroregions of Italy). The pandemic initial years of 2020 and 2021 presented the lowest reporting. While disruption to health services and awareness of infections beyond COVID-19 may have faltered during this period, we also uncovered that such years presented some of the lowest climate-based transmission suitability. Higher reporting was observed for 2019 and 2023, which did present higher than normal suitability, but that more likely reflected the success of dengue epidemic activity elsewhere. For example, 2019 was a year of resurgence in South America and 2023 is now a record year for incidence across the globe. Over the years, three of the four dengue serotypes have been reported to circulate in Italy. Nonetheless, their success in being introduced into the country should also reflect epidemic expansion elsewhere, specifically in light of the absence of herd-immunity in Italy.

The year of 2023 in Italy was exceptional, not just due to the total number of reported cases, but also for the uncommon co-occurrence of a few autochthonous transmission chains of more than one serotype. We summarize that such chains occurred mostly within two spatial clusters, one in the Northern and another in the Center macroregions. Age profiles for 2023 were available and revealed a higher incidence in the 20-39 age group. The latter contrasted with the national age pyramid of the country, suggesting that active age individuals were more exposed to infection. This effect was particularly strong for females, while males presented a more uniform incidence above 20 years of age. The reasons for this asymmetric exposure are unknown and should be the focus of future epidemiological studies.

Despite the limited number of available dengue genomes related to infections reported in Italy, we were able to recover a few observations of interest. We noted, for example, that viral lineages of the 2023 transmission clusters belonged to DENV1 genotype V and DENV3 genotype III. Notably, circulation of DENV1 resulted from two independent introductions into the country, while DENV3 resulted from a single introduction. The origin of DENV1 was suggested to be from South America, while the origin of DENV3 is likely to have been the Caribbean. Once again, we noted that the co-circulation of DENV1 and DENV3 in Italy in 2023 conspicuously occurred while the specific genotypes experienced large expansion in Latin American countries [20, 21]. It should be noted however, that the number of genomes available limit definite conclusions, and as such the results presented in this study are amenable to change upon inclusion of further genomic data.

The information summarized in this study highlights the necessity of a strong genomic surveillance network in Italy for tracking and identifying introductions and local chains of transmission, primarily facilitated by the movement of viremic travelers from elsewhere. A shift from passive to active surveillance would improve responsiveness to short-term transmission chains similar to those experienced during 2023. In parallel, active surveillance strategies should take into account factors such as the spatio-temporal variation of climate suitable for local transmission and human mobility from endemic countries. Together, this could help ensure a comprehensive approach to monitoring and containing dengue introductions and arboviruses more generally.

## Conflict of Interest

The authors declare that there are no conflicts of interest.

## Author Contribution

**Conception and design:** F.B., M.C., J.L., M.G.; **Investigations:** F.B., T.N., M.G., J.L; Data Analysis: F.B., T.N., J.L., M.G.; **Visualization:** F.B., J.L., M.G.; **Writing – Original:** F.B., J.L., M.G.; **Writing – Revision**: F.B., A.M., T.N., F.S., A.C., L.C.J.A., A.M.B.d.F., A.M., J.L., M.C., M.G.

## Acknowledgments

M. Giovanetti’s funding is provided by PON “Ricerca e Innovazione” 2014-2020 and by the CRP-ICGEB RESEARCH GRANT 2020 Project CRP/BRA20-03, Contract CRP/20/03.

## Data Availability

The data that support the findings of this study are openly available at the following link: https://github.com/fbranda/arbovirus.

---

**Table S1.** Overview of File Structure for the Italian Dengue virus Data Repository.

| Files and format | Description |
| --- | --- |
| **Bulletins**<br>PDF | These files provide updates on the Dengue situation throughout Italy |
| **Dengue-ita-yyyy\*.csv**<br>Comma-separated values file (.csv, UTF-8 encoded) | Number of Reported Dengue Cases Nationwide, Categorized by Sex and Type of Infection (Autochthonous and Imported) |
| **Dengue-ita-age-yyyy\*.csv**<br>Comma-separated values file (.csv, UTF-8 encoded) | Number of Dengue Cases Reported by Age and Sex |
| **Dengue-ita-location-exposure-yyyy\*.csv**<br>Comma-separated values file (.csv, UTF-8 encoded) | Percentage of Dengue Cases Relative to Total Cases by Location |
| **Dengue-ita-regions-yyyy\*.csv**<br>Comma-separated values file (.csv, UTF-8 encoded) | Reported Dengue Cases by Region |
| **Dengue-ita-summary-cases.csv**<br>Comma-separated values file (.csv, UTF-8 encoded) | Time Series of Nationwide Reported Dengue Cases by Year, Month, and Type of Infection (Autochthonous and Imported) |
| **Dengue-ita-summary-cases-regions.csv**<br>Comma-separated values file (.csv, UTF-8 encoded) | Time Series of Regionally Reported Dengue Cases Categorized by Year, Month, and Type of Infection (Autochthonous and Imported) |

\*yyyy is the year of the monitoring

## Notes

### Competing Interest Statement

The authors have declared no competing interest.

### Funding Statement

Not Applicable

